# Analyzing greedy vaccine allocation algorithms for metapopulation disease models

**DOI:** 10.1101/2024.10.12.24315394

**Authors:** Jeffrey Keithley, Akash Choudhuri, Bijaya Adhikari, Sriram V. Pemmaraju

## Abstract

As observed in the case of COVID-19, effective vaccines for an emerging pandemic tend to be in limited supply initially and must be allocated strategically. The allocation of vaccines can be modeled as a discrete optimization problem that prior research has shown to be computationally difficult (i.e., NP-hard) to solve even approximately.

Using a combination of theoretical and experimental results, we show that this hardness result may be circumvented. We present our results in the context of a metapopulation model, which views a population as composed of geographically dispersed heterogeneous subpopulations, with arbitrary travel patterns between them. In this setting, vaccine bundles are allocated at a subpopulation level, and so the vaccine allocation problem can be formulated as a problem of maximizing an integer lattice function 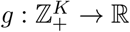 subject to a budget constraint ∥**x**∥_1_ ≤ *D*. We consider a variety of simple, well-known greedy algorithms for this problem and show the effectiveness of these algorithms for three problem instances at different scales: New Hampshire (10 counties, population 1.4 million), Iowa (99 counties, population 3.2 million), and Texas (254 counties, population 30.03 million). We provide a theoretical explanation for this effectiveness by showing that the approximation factor of these algorithms depends on the *submodularity ratio* of objective function *g*, a measure of how distant *g* is from being submodular.

**Author summary:** Strategic and timely allocation of vaccines is crucial in combating epidemic outbreaks. Developing strategies to allocate vaccines over sub-populations rather than to individuals leads to policy recommendations that are more feasible in practice. Despite this, vaccine allocation over sub-populations has only received limited research interest, and the associated computational challenges are relatively unknown. To address this gap, we study vaccine allocation problems over geographically distinct subpopulations in this paper. We formulate our problems to reduce either **i)** the total infections or **ii)** the sum of peak infections over meta-population disease models. We first demonstrate that these problems are computationally challenging even to approximate and then show that a family of simple, well-known greedy algorithms exhibit provable guarantees. We conduct realistic experiments on state-level mobility networks derived from real-world data in three states of distinct population levels: New Hampshire, Iowa, and Texas. Our results show that the greedy algorithms we consider are **i)** scalable and **ii)** outperform both state-of-the-art and natural baselines in a majority of settings.

## Introduction

In the early stages of a pandemic like COVID-19, the demand for vaccinations far exceeds supply [1, 2] and it is critical to strategically allocate vaccines [3, 4]. The vaccine allocation problem can be modeled in a variety of ways, including as discrete optimization problems [5–9]. However, all of these problems are computationally hard, even to solve approximately (see [10], for a specific example). Despite these obstacles, we need to be able to solve vaccine allocation problems at scale and have confidence that the obtained solutions are close to being optimal. In this paper, we take steps towards this goal.

We consider the metapopulation-network model for disease-spread [11, 12], which allows for heterogeneity among geographically distinct subpopulations and arbitrary travel patterns between them. Vaccine allocation within this model consists of allocating some number of bundles of vaccines to each subpopulation while satisfying an overall budget constraint. The resulting family of problems, which we call the *Metapopulation Vaccine Allocation (MVA)* problems, can be formalized as maximizing an objective function (e.g., number of cases averted) defined over an integer lattice domain subject to a budget constraint. Not surprisingly, we show specific problems obtained via realistic instantiations of the metapopulation-network model and objective function in MVA are not just NP-hard, but even hard to approximate. We show these hardness results for two instantiations, which we call MaxCasesAverted and MaxPeaksReduced, of MVA over SEIR (**S**usceptible-**E**xposed-**I**nfected-**R**ecovered) metapopulation models [11, 12].

These hardness of approximation results imply that worst-case approximation guarantees are not attainable for natural instantiations of MVA. However, for a family of simple, well-known greedy algorithms, we show positive theoretical and experimental results for both MaxCasesAverted and MaxPeaksReduced. These simple and natural greedy algorithms lend themselves to the machinery from submodular function optimization for in-depth analysis. There is a rich literature of methods for submodular set function optimization [13–18] that has subsequently been extended to submodular optimization over the integer lattice [19–22]. Furthermore, in the last few years, researchers have attempted to extend some of the aforementioned results for submodular set and lattice function optimization to functions that are not submodular, by using the notion of *submodularity ratio* of a function, which is a measure of how distant that function is from being submodular [23–25]. All of this literature is foundational to our approach to analyzing vaccine allocation algorithms in a metapopulation model setting [11, 12].

In our main theoretical result, we show that simple greedy algorithms provide worst-case approximation guarantees for MaxCasesAverted and MaxPeaksReduced that become better as the *submodularity ratio* of their objective functions approaches 1. The submodularity ratio [23–26] of a set or lattice function is a measure (between 0 and 1) of how close the function is to being submodular, with values closer to 1 corresponding to functions that are closer to being submodular. We complement this theoretical result with experimental results indicating that the objective functions for MaxCasesAverted and MaxPeaksReduced might have relatively high submodularity ratios.

We then experimentally evaluate the performance of greedy vaccine allocation algorithms at three scales; we use New Hampshire (10 counties, population 1.4 million) for our small scale experiments, Iowa (99 counties, population 3.2 million) for our medium scale experiments, and Texas (254 counties, population 30.03 million) for our large scale experiments. We compare the performance of the greedy methods with a set of trivial baselines, such as allocating vaccines according to population sizes. We also compare against a randomized algorithm called Pareto Optimization for Subset Selection (POMS) [24]. POMS works by expanding a random pareto-optimal frontier, and was designed to compete against greedy algorithms for small scale problems. We show the greedy algorithms we consider outperform POMS for our experimental settings, while scaling more readily. Our experiments demonstrate that **i)** simple greedy vaccine allocation algorithms outperform the natural baseline algorithms substantially (up to 9M more individuals saved than the worst-performing baseline in some settings), **ii)** for both MaxCasesAverted and MaxPeaksReduced, greedy algorithms perform near-optimally for most problem instances we evaluate for New Hampshire (and recover similar approximation guarantees to those of submodular functions for experiments in Iowa and Texas), and **iii)** the fastest of our greedy algorithms are feasible even for large scale instances such as the state of Texas.

## Materials and methods

### Background

#### Notation

We use ℤ_+_ to denote the set of non-negative integers and for any positive integer *n*, we use [*n*] to denote the set {1, 2, …, *n*}.

#### Metapopulation disease-spread models

A *metapopulation* disease-spread model [11] generalizes the classic homogeneous-mixing compartmental models [27], by allowing geographically-diverse subpopulations. Let *K* ∈ ℤ_+_ denote the number of subpopulations in the metapopulation model. For each subpopulation *i* ∈ [*K*], let *n*_*i*_ denote the size of the subpopulation and let **n** denote the vector (*n*_1_, *n*_2_, …, *n*_*K*_) of subpopulation sizes. For each pair (*i, j*) ∈ [*K*] × [*K*], let *w*_*ij*_ ∈ ℤ_+_ denote the number of individuals moving from subpopulation *i* to subpopulation *j* daily. Thus, each *w*_*ij*_ is a static (i.e., time independent) quantity. Let **W** denote the *K* × *K* mobility matrix induced by the *w*_*ij*_ values.

Our goal is to decrease the spread of disease by allocating a total of *D* ∈ ℤ_+_ bundles of vaccines to individuals over all subpopulations; here *D* is the *vaccine budget*. A bundle can be viewed as the smallest “shipment” of vaccines that can be allocated to a subpopulation and we assume that each bundle consists of an integer Δ *>* 0 number of individual vaccines. Let 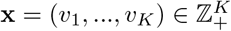 denote a vaccine allocation, where *v*_*i*_ is the number of bundles of vaccines allocated to subpopulation *i*. For simplicity, we assume that vaccination is preemptive, i.e., occurs at time 1, with knowledge of initial infected, but before the disease has started to spread. It is straightforward to generalize this to a setting in which vaccine allocation occurs later in the progression of the disease. Let 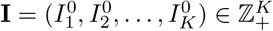,where 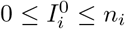, denote the number of initial infections in subpopulation *i*. Let *f* (**x** | ℳ, **I**) denote some measure of disease-spread according to the metapopulation model ℳ starting with initial infection vector **I**, expressed as a function of the vaccine allocation vector **x**. For example, *f* (**x** | ℳ, **I**) could denote the total number of infected individuals over some time window. Let *g*(**x** | ℳ, **I**) denote *f* (**0** | ℳ, **I**) − *f* (**x** | ℳ, **I**), representing the reduction in disease-spread due to vaccine allocation 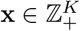,relative to the no-vaccine setting. Note that both *f* and *g* are defined over the integer lattice 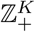 and our goal is to maximize the integer lattice function *g*(**x** | ℳ, **I**) subject to the cardinality constraint ∥**x**∥_1_ ≤ *D*. **Submodularity of lattice functions** For *K* ∈ ℤ_+_, let 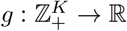 be a function defined on an integer lattice domain. The function *g* is said to be *submodular* if for all **x, y** ∈ ℤ_+_

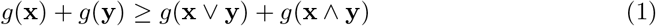

Here (**x** ∨ **y**)_*i*_ = max{**x**_*i*_, **y**_*i*_} and (**x** ∧ **y**)_*i*_ = min{**x**_*i*_, **y**_*i*_}.

Below we provide an alternate “diminishing returns” notion of submodularity that is easier to work with. Here **e**_*i*_ denotes the unit vector with 1 in coordinate *i*.

##### Definition 1

*[21]* ***(DR-Submodularity)*** *A function* 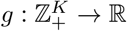 *is said to be diminishing returns submodular (DR-submodular, in short) if g*(**x** + **e**_*i*_) − *g*(**x**) ≥ *g*(**y** + **e**_*i*_) − *g*(**y**) *for all i* ∈ [*K*] *and* 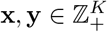, *where* **x** ≤ **y**.

For set functions, submodularity and DR-submodularity are equivalent. However, it is known [20] that if a lattice function is DR-submodular then it is submodular, but the converse is false. Thus, DR-submodularity is a stronger notion compared to submodularity. However, [24] presents a DR-type characterization of submodular lattice functions that is quite useful for our analysis.

##### Lemma 2.

*[24] A function* 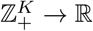 *is submodular if and only if for any* 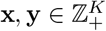, **x** ≤ ***y*** *and i* ∈ [*K*] *with* **x**_*i*_ = **y**_*i*_, *g*(**x** + **e**_*i*_) − *g*(**x**) ≥ *g*(**y** + **e**_*i*_) − *g*(**y**).

Note that according to this lemma, for submodular lattice functions, the DR property is only required to hold at identical coordinates of **x** and **y**.

The computational complexity of maximizing a submodular lattice function 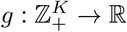 subject to a cardinality constraint, namely max_∥**x**∥ ≤*D*_ *g*(**x**), is well understood. [20] extend the result for set functions from [28] to lattice functions and show that greedy approaches yield a 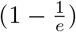 -approximation for this problem for both submodular and DR-submodular lattice functions. These approximation guarantees are optimal due to the inapproximability result of [29].

### The SEIR Metapopulation model

The SEIR equations are governed by parameters *λ, η*, and *δ*, where *λ* is the *infectivity*, 1*/η* is the *latency period*, and 1*/δ* is the *infectious period*. Let *r*_*i*_ denote a multiplier that scales *λ* to allow for county differences in contact rates. Let *T* be a positive integer denoting the size of the time window under consideration. For *t* ∈ [*T*] ∪ {0}, each subpopulation is split into compartments 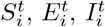 and 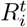 representing the number of susceptible, exposed, infected, and recovered individuals within subpopulation *i* at time *t*. We assume the initial conditions 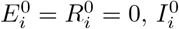 is an arbitrary non-negative number satisfying 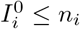, and 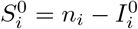. The evolution of 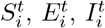,and 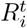 over time *t* is respectively governed by equations (2)-(5). The term 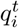 that appears in these equations is called the *force of infection*. When 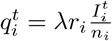 equations (2)-(5) represent the spread of disease in a single subpopulation *i* with a homogeneous mixing assumption.

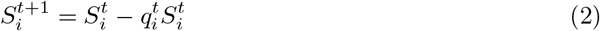

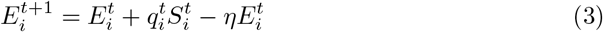

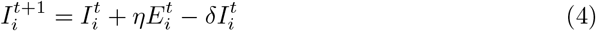

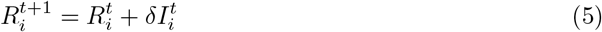

We use the following expression for the force of infection term 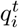 that takes the infection incidence within subpopulation *i* along with flows of individuals into and out of subpopulation *i*. The derivation of 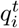 is inspired by a similar derivation in [5, 12] and is included in the Supplementary Information.

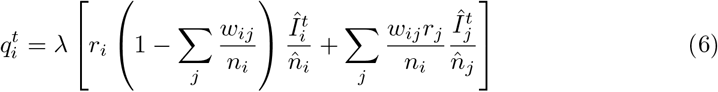

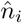 denotes the *effective* population of subpopulation *i* at time *t*, describing the number of individuals present in subpopulation *i* after a daily commute has occurred, and 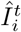 denotes the effective number of infected individuals in subpopulation *i* after a commute. The first term in the right hand side of Eqn (6) is the proportion of individuals leaving subpopulation *i* for their commute, and the second term is the proportion of individuals arriving.

The SEIR metapopulation model ℳ described above is completely specified by the vector (**n, r, W**, *T, λ, η, δ*). In our experiments, each subpopulation represents a county within a state (e.g., *K* = 99 for Iowa) and the mobility matrix **W** is obtained from two independent sources, FRED [30] and SafeGraph [31]. By instantiating a specific disease-spread model for each subpopulation and describing its interaction with mobility matrix **W**, we can obtain a completely specified metapopulation model.

Table 1 summarizes the notation introduced in this section.

**Table 1.** Metapopulation model notation.

| Variable | Definition |
| --- | --- |
| $K, T$ | Number of subpopulations, size of time window |
| $r_i$ | Population density correlated λ-multiplier for subpopulation $i$ |
| $n_i$ | Size of subpopulation $i$ |
| $w_{ij}$ | Mobility from subpopulations $i$ to $j$ |
| $q_i^t$ | Force of infection in subpopulation $i$ at time $t$ |
| $\lambda, 1/\eta, 1/\delta$ | Infectivity, latency period, infectious period |

**Table 2.**
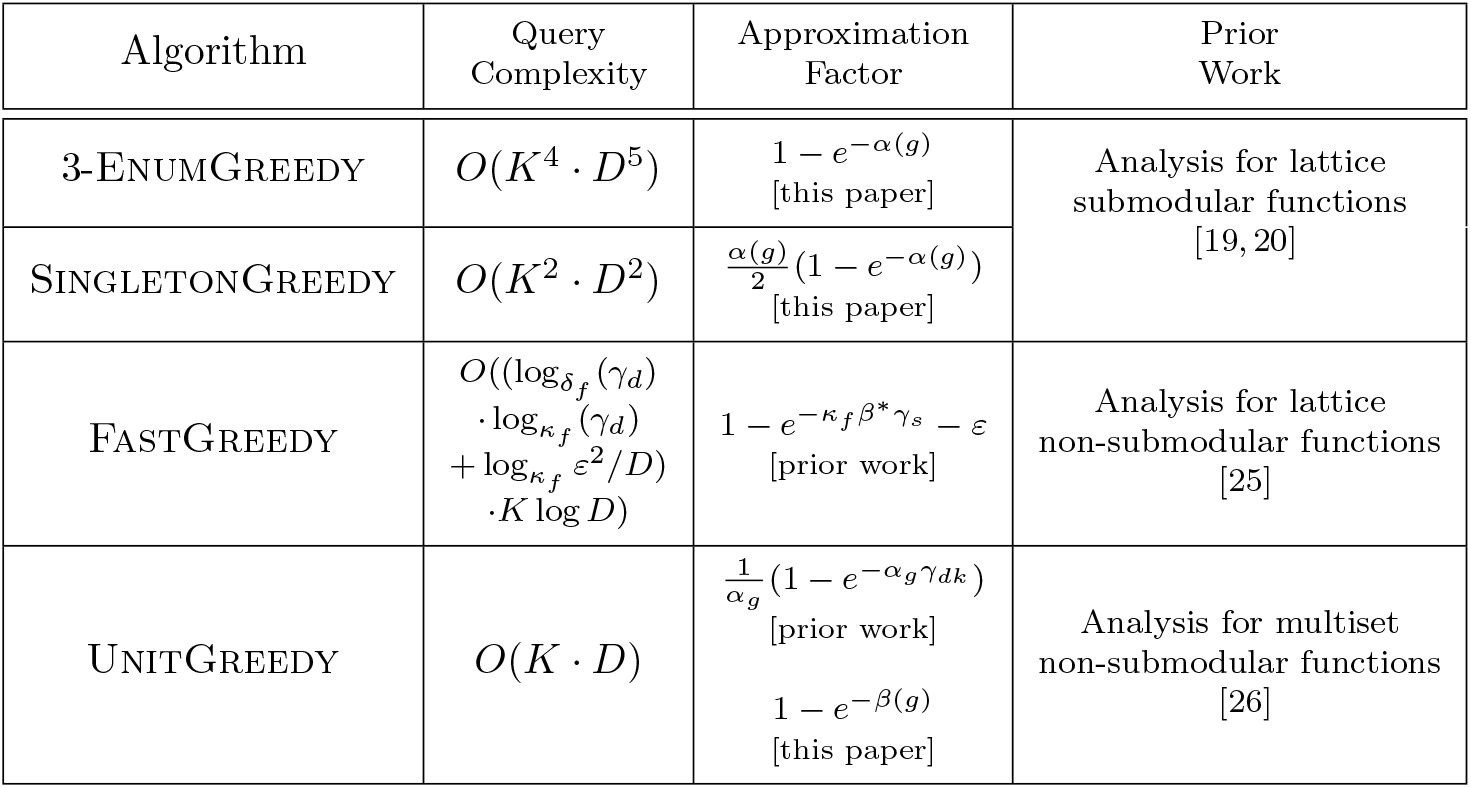
Summary of greedy algorithms presented in this section. Details of approximation factors for FastGreedy and UnitGreedy may be found in [26] and [25], respectively.

| Algorithm | Query Complexity | Approximation Factor | Prior Work |
| --- | --- | --- | --- |
| 3-ENUMGREEDY | $O(K^4 \cdot D^5)$ | $1 - e^{-\alpha(g)}$<br>[this paper] | Analysis for lattice submodular functions<br>[19, 20] |
| SINGLETONGREEDY | $O(K^2 \cdot D^2)$ | $\frac{\alpha(g)}{2}(1 - e^{-\alpha(g)})$<br>[this paper] | |
| FASTGREEDY | $O((\log_{\delta_f}(\gamma_d) \cdot \log_{\kappa_f}(\gamma_d) + \log_{\kappa_f} \varepsilon^2/D) \cdot K \log D)$ | $1 - e^{-\kappa_f \beta^* \gamma_s} - \varepsilon$<br>[prior work] | Analysis for lattice non-submodular functions<br>[25] |
| UNITGREEDY | $O(K \cdot D)$ | $\frac{1}{\alpha_g}(1 - e^{-\alpha_g \gamma_{dk}})$<br>[prior work]<br>$1 - e^{-\beta(g)}$<br>[this paper] | Analysis for multiset non-submodular functions<br>[26] |

### Problem formulations

We are now ready to state the *Metapopulation Vaccine Allocation* (MVA) family of problems.

#### MVA

Given a metapopulation model ℳ, initial infected vector 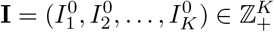, where 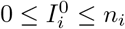,and a vaccine budget *D* ∈ ℤ_+_, find a vaccine allocation 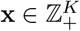, satisfying ∥**x**∥_1_ ≤ *D* such that *g*(**x** | ℳ, **I**) := *f* (**0** | ℳ, **I**) − *f* (**x** | ℳ, **I**) is maximized.

### SEIR Metapopulation Vaccine Allocation Problems

For illustrative purposes, we instantiate the general metapopulation model ℳ with an SEIR model for disease spread within each subpopulation. Our framework is general and the SEIR model that we use within subpopulations can be replaced by any other homogeneous-mixing disease spread model.

Using the SEIR metapopulation model described above, we obtain specific instances of the MVA problem. But before we can describe these specific instances, we need to describe how vaccination affects disease spread in the SEIR metapopulation model. For simplicity, we assume that vaccine uptake and vaccine effectiveness are both perfect, and thus allocating a vaccine bundle **x** = (*v*_1_, …, *v*_*K*_) implies that Δ · *v*_*i*_ individuals in subpopulation *i* are vaccinated and removed from 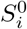.Thus the vaccine allocation **x** = (*v*_1_, …, *v*_*K*_) updates the initial susceptible to 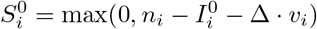 for all *i* ∈ [*K*]. The assumptions of perfect uptake and effectiveness are easily relaxed; lowering the vaccine uptake or effectiveness is equivalent to allocating fewer vaccines.

We now present two illustrative problems that maximize the impact of vaccines according to different disease spread metrics. In the problem MaxCasesAverted, the metric is the total number of infections averted across all subpopulations, and in the problem MaxPeaksReduced, the metric is the decrease in the sum of all infection peaks across all subpopulations (both taken over the entire simulation time). More precisely, given an SEIR metapopulation model ℳ = (**n, r, W**, *T, λ, η, δ*), initial infected vector 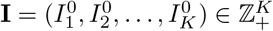,, where 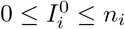,and a vaccine allocation 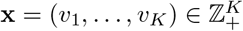,we define the metric

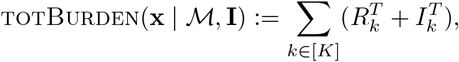

which is simply the total number of individuals who became infected in the time window [0, *T*]. Another natural disease spread metric for the SEIR metapopulation model is

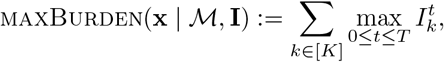

which is the total number of individuals infected during “peak” infection time over all the subpopulations. This metric is motivated by the fact that even small peaks are challenging in low-resource counties (typically in low-population counties), because healthcare infrastructure is often limited in such counties. So even a small spike in the number of infected individuals can quickly overwhelm local resources. Thus we seek to reduce the likelihood that local healthcare systems will be overwhelmed with the maxburden metric. Given metapopulation model ℳ, initial infection vector **I**, and budget *D*, we define the following discrete optimization problems:

#### MaxCasesAverted

Find a vaccine allocation 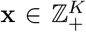, satisfying ∥**x**∥_1_ ≤ *D* such that the following is maximized.

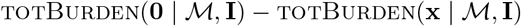

#### MaxPeaksReduced

Find a vaccine allocation 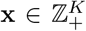,satisfying ∥**x**∥_1_ ≤ *D* such that the following is maximized.

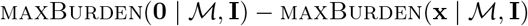

### Hardness of MaxCasesAverted and MaxPeaksReduced

As with many resource allocation problems, both MaxCasesAverted and MaxPeaksReduced are not only NP-hard, but even hard to efficiently approximate. We show this by a reduction from the *Maximum k-Subset Intersection* (maxksi) problem [32]. The input to Max *k*-SI consists of a collection 𝒞 = {*S*_1_, *S*_2_, …, *S*_*m*_} of sets, where each set *S*_*j*_ is a subset of a universe 𝒰 = {*e*_1_, *e*_2_, …, *e*_*n*_}, and a positive integer *k*. The problem seeks to find *k* subsets 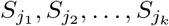 from 𝒞 whose intersection has maximum size. The following theorem from [32] shows that Max *k*-SI is highly unlikely to have an efficient approximation algorithm, even with an inverse polynomial approximation factor.

#### Theorem 3.

*[32] Let ϵ >* 0 *be an arbitrarily small constant. Assume that SATISFIABILITY does not have a probabilistic algorithm that decides whether a given instance of size n is satisfiable in time* 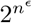. *Then there is no polynomial time algorithm for* Max *k*-SI *that achieves an approximation ratio of* 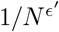, *where N is the size of the given instance of* Max *k*-SI *and ϵ*^*′*^ *only depends only on ϵ*.

We now show a reduction from Max *k*-SI to both MaxCasesAverted and MaxPeaksReduced, thereby establishing the inapproximability of both of these problems.

#### Theorem 4.

*Let ϵ >* 0 *be an arbitrarily small constant. Assume that SATISFIABILITY does not have a probabilistic algorithm that decides whether a given instance of size n is satisfiable in time* 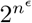. *Then there is no polynomial time algorithm for* MaxCasesAverted *or for* MaxPeaksReduced *that achieves an approximation ratio of*1*/N* 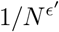, *where N is the size of the given instance of* MaxCasesAverted *or* MaxPeaksReduced *and ϵ*^*′*^ *only depends only on ϵ*.

**Proof:** To prove the portion of this theorem pertaining to MaxCasesAverted, we show the following lemma.

#### Lemma 5.

*Suppose there is a polynomial-time algorithm* 𝒜 *that yields an α-approximation for* MaxCasesAverted. *Then there is a polynomial-time α/*2*-approximation algorithm* 𝒜^*′*^ *for* Max *k*-SI.

**Proof of Lemma 5**. Given an instance (𝒞, 𝒰, *k*) of Max *k*-SI, we construct the graph *G* with *m* + *n* + 1 nodes. For each subset *S*_*j*_ ∈ 𝒞 and each *e*_*i*_ ∈ 𝒰, there is a node in *G*, for a total of *m* + *n* nodes. There is an extra node *I* that is connected to every *S*_*j*_-node. There are edges between the *S*_*j*_-nodes and the *e*_*i*_-nodes connecting an *S*_*j*_-node to an *e*_*i*_-node iff *e*_*i*_ ∉ *S*_*j*_.

To each node *v* in *G*, we assign a population *n*_*v*_ as follows: 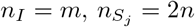 for all *j* ∈ [*n*], and 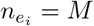 for all *i* ∈ [*n*], where *M* is a large integer whose value will be specified later. We then interpret each undirected edge in *G* as a pair of directed edges pointing in opposite directions and assign a flow to each directed edge. We assign flow 1 to each edge from *I* to *S*_*j*_ and to each edge from *S*_*j*_ to *e*_*i*_. To all other edges, i.e., the edges pointing “backwards”, we assign flow 0. This construction is illustrated in Fig 1. This specifies the vectors **n** and **w** of the instance of MaxCasesAverted.

**Fig 1.**
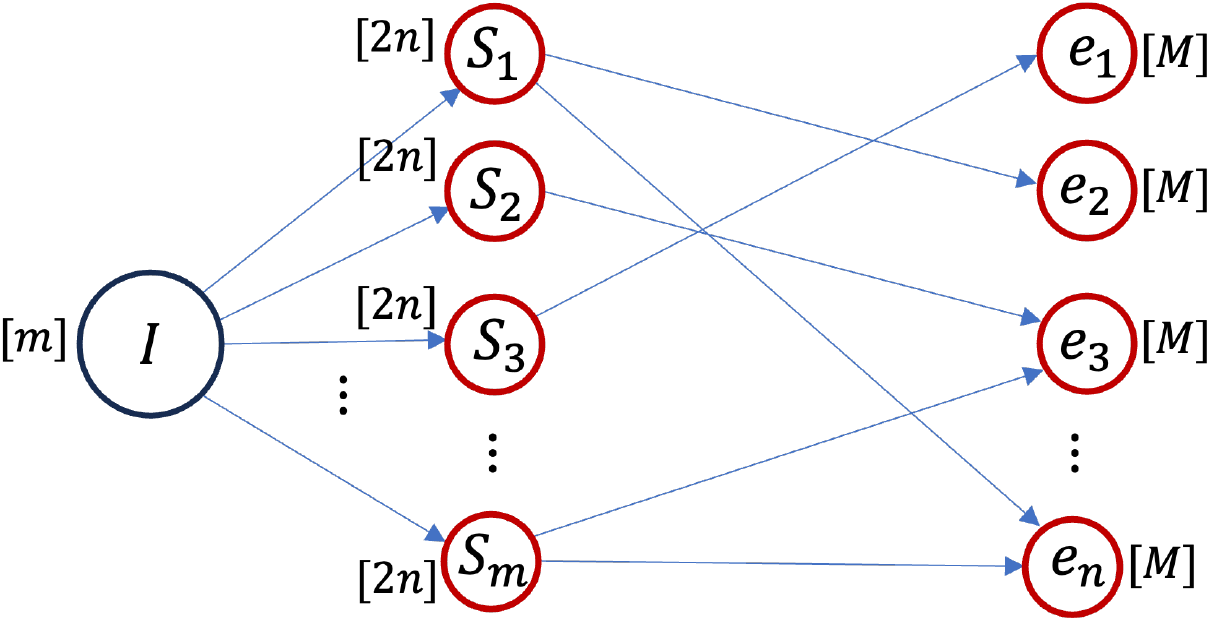
The instance of MaxCasesAverted and MaxPeaksReduced is a graph *G* constructed from the given instance (𝒞 = (*S*_1_, *S*_2_, …, *S*_*m*_), 𝒰 = (*e*_1_, *e*_2_, …, *e*_*n*_), *k*) of Max *k*-SI. Each node represents a subpopulation, with the size of the subpopulation shown in square brackets next to it. The directed edges permit 1 unit flow. The unit flows from nodes *S*_*j*_ to *e*_*i*_ encode non-membership. For example, the flow from *S*_1_ to *e*_2_ implies that *e*_2_ ∉ *S*_1_.

We set the contact rate *r*_*v*_ and infectivity *λ* such that the force of infection 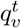 is always at least 1. This corresponds to “perfect infectivity”, meaning that if a subpopulation contains some infected and some susceptible individuals at a time step, then *all* the susceptible individuals in the subpopulation will transition to the exposed state at the next time step. We then set *η* = *δ* = 1 so that the latency period and recovery period are both 1. With this setting of the parameters, the infection will completely die out in 5 time steps, i.e., every individual will either be susceptible or recovered. So we set the size of the time window *T* = 5. Finally, we set the vaccination budget *D* = (*m* − *k*) · 2*n* and initialize the entire population of *m* individuals at node *I* to be infected and all other individuals to be susceptible. This completes the specification of the problem instance ℐ of MaxCasesAverted.

We now make 2 simple observations that follow from the construction of ℐ and depend on the notion of being “unprotected” with respect to a vaccine allocation. Let **x** be an arbitrary, feasible allocation for ℐ. A subpopulation *S*_*j*_ is called *unprotected for* **x** if 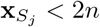 ;otherwise, *S*_*j*_ is called *protected for* **x**. A subpopulation *e*_*i*_ is called *unprotected for* **x** if 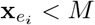 *M* and for some subpopulation *S*_*j*_ that is unprotected for **x**, the edge {*S*_*j*_, *e*_*i*_} is in *G*; otherwise, *e*_*i*_ is called *protected for* **x**.

#### Observation 1

In every unprotected subpopulation 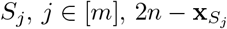,individuals will become exposed in time step 1 and infected in time step 2.

#### Observation 2

In every unprotected subpopulation 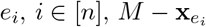,individuals will become exposed in time step 3 and infected in time step 4.

These 2 observations immediately lead to the following 3 claims.

#### Claim i)

Consider a vaccine allocation 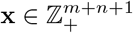 that is feasible for ℐ and satisfies 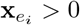. Let 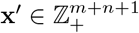 be an allocation obtained from **x** by reallocating all vaccines from the subpopulation *e*_*i*_ to subpopulations *S*_*j*_, *j* ∈ [*m*]. Then **x**^*′*^ is feasible for ℐ and

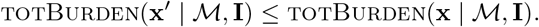

#### Claim ii)

Consider a vaccine allocation 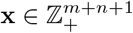 that is feasible for ℐ and satisfies 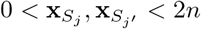 for two subpopulations *S*_*j*_, *S*_*j*′_, *j* ≠ *j*^*′*^. Let 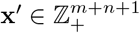 be an allocation obtained from **x** by reallocating as many vaccines as possible from the subpopulation *S*_*j*_^*′*^ to the subpopulation *S*_*j*_, until 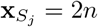 or 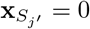 (or both). Then **x**^*′*^ is feasible for ℐ and

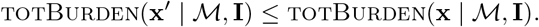

#### Claim iii)

Consider a vaccine allocation 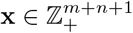 that is feasible for ℐ and satisfies ||*x*||_1_ = *D* = (*m* − *k*) · 2*n*. Then using the reallocations from Claims (i) and (ii), it is possible to transform **x** into 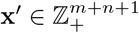 in polynomial time such that **x**^*′*^ is feasible for ℐ,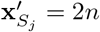 for exactly (*m* − *k*) subpopulations *S*_*j*_, **x**^*′*^ is 0 for all other subpopulations, and

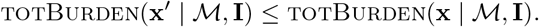

Claim (iii) allows us to assume that any *α*-approximation algorithm 𝒜 for MaxCasesAverted returns an allocation **x**^*′*^ for the problem instance ℐ, that picks exactly (*m* − *k*) subpopulations *S*_*j*_ and vaccinates these subpopulations entirely, while allocating no vaccines to any of the remaining subpopulations. Similarly, Claim (iii) implies that there is an optimal allocation **x**^*^ for ℐ that picks exactly (*m* − *k*) subpopulations *S*_*j*_ and vaccinates these subpopulations entirely, while allocating no vaccines to any of the remaining subpopulations.

Let 𝒮(**x**^*′*^) be the set of subpopulations *S*_*j*_ unprotected for **x**^*′*^. Similarly, define 𝒮(**x**^*^). Note that |𝒮(**x**^*′*^)| = |𝒮(**x**^*^)| = *k*. Let ℰ(**x**^*′*^) be the set of subpopulations *e*_*i*_ that are protected for **x**^*′*^. Similarly, define ℰ(**x**^*^). By the construction of edges from subpopulations *S*_*j*_ to subpopulations *e*_*i*_ in ℐ, it follows that 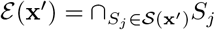.Similarly, 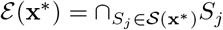

The objective function value of MaxCasesAverted for the optimal allocation **x**^*^, which is totburden(**0** | ℳ, **I**) − TotBurden(**x**^*^ | ℳ, **I**), can be simplified to

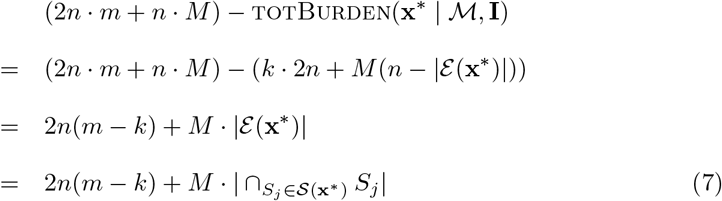

Similarly, the objective function value of MaxCasesAverted for the *α*-approximate allocation **x**^*′*^ is 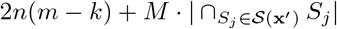 Since **x**^*^ maximizes the objective function value of MaxCasesAverted, Equation (7) implies that 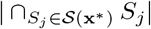 has largest possible cardinality. Since |𝒮(**x**^*^)| = *k*, this implies that 𝒮(**x**^*^) is an optimal solution to the Max *k*-SI problem. Using *OPT*_Max*k*-SI_ to denote the optimal objective function value of Max *k*-SI, we can rewrite the expression (7) as 2*n*(*m* − *k*) + *M* · *OPT*_Max *k*-SI_. Since **x**^*′*^ is an *α*-approximate solution to MaxCasesAverted,

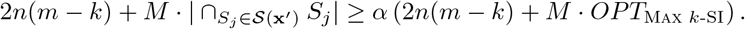

Rearranging terms we get

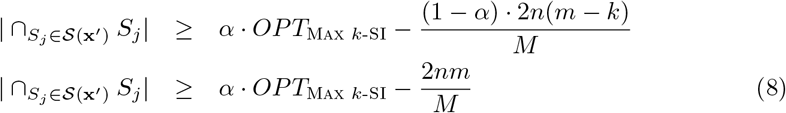

Picking *M* large enough so that 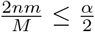 and using *OPT*_Max *k*-SI_ ≥ 1, we obtain

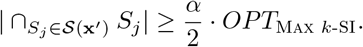

This implies that the allocation **x**^*′*^ can be used to obtain an *α/*2-approximation to Max *k*-SI.

We now prove a similar lemma for the MaxPeaksReduced problem.

#### Lemma 6.

*Suppose there is a polynomial-time algorithm* 𝒜 *that yields an α-approximation for* MaxPeaksReduced. *Then there is a polynomial-time α-approximation algorithm* 𝒜^*′*^ *for* Max *k*-SI.

**Proof of Lemma 6**. This uses the same argument as the lemma above. Claims (i) and (ii) hold for MaxBurden(**x** | ℳ, **I**) as well and from these two claims, Claim (iii) follows. Furthermore,

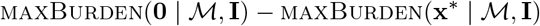

simplifies exactly to expression (7), from which inequality (8) follows. From this, the lemma immediately follows, as shown above.

### Algorithmic approach and analysis

We consider a variety of greedy algorithms for MVA. These algorithms and their accompanying analyses also apply to the general budget-constrained maximization problem on an integer lattice: 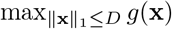 where 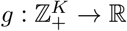 is an arbitrary, monotone function defined on an integer lattice. We start by defining the LatticeGreedySubroutine, whose search space is the entire lattice ℤ^*K*^ in each iteration. This subroutine forms the basis for two greedy algorithms *ℓ*-EnumGreedy and SingletonGreedy [20] (detailed below). Algorithms based on LatticeGreedySubroutine are prohibitively expensive for large problem instances, so we also consider FastGreedy [25], which is a relaxation of LatticeGreedySubroutine, based on a threshold greedy algorithm [33]. In addition, we evaluate and further analyze an approach which considers the lattice as a multiset and runs the greedy algorithm for set functions over it, which we call UnitGreedy (Algorithm 3). In this section, we describe each algorithm we evaluate and their associated approximation guarantees, some of which we derive.

### Greedy algorithm descriptions

#### LatticeGreedySubroutine Description

As shown in the Algorithm 1 pseudocode, LatticeGreedySubroutine selects a (*k*^*^, *s*^*^) pair that maximizes the marginal gain of *g*(·) in each iteration, where *k*^*^ ∈ [*K*] is a subpopulation and *s*^*^ ∈ ℤ_+_ is the number of bundles to allocate to subpopulation *k*^*^. To compute the highest marginal gain among all possible (*k, s*) ∈ [*K*] × ℤ_+_ pairs in each iteration of the algorithm, we assume that the algorithm has access to a “value oracle” that returns the value of the objective function *g*(·) at any point in its domain. It is possible that the selected pair (*k*^*^, *s*^*^) is not feasible because adding it to the solution causes the budget constraint to be violated. Such an iteration is said to have *failed*, and we remove the (*k*^*^, *s*^*^) pair from the search space *Q*. Otherwise, the iteration is *successful* and the (*k*^*^, *s*^*^) pair is used to update the allocation. It is useful for our analysis to state the algorithm in this manner, allowing for failed iterations. However, to obtain an efficient implementation we can, in Line 4, prune the search space *Q* so as to guarantee that the condition in Line 5 is always satisfied. Such an implementation runs in *O*(*K* · *D*^2^ · *T*_*g*_) time in the worst case, where *T*_*g*_ is the worst case running time of the value oracle. However, the at most *K* · *D* pairs in *Q* can all be evaluated in parallel, and assuming full parallelism with no overhead, the running time of LatticeGreedySubroutine can also be reduced to *O*(*D* · *T*_*g*_ · log(*K* · *D*)) in the PRAM model (even with exclusive read and exclusive write). We note that LatticeGreedySubroutine and the algorithms based on it come from [20].

We further allow LatticeGreedySubroutine to start with an arbitrary initial allocation 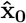,and not just **0** (see Line 1). This is so that we can use LatticeGreedySubroutine as the completion step for an algorithm that enumerates solutions of bounded size. Specifically, let *ℓ* ≥ 1 be a fixed integer and let 𝒮 be the set of all feasible solutions of size *ℓ* or less. Thus each element in 𝒮 is a subset of at most *ℓ* subpopulations, each allocated some number of vaccine bundles so that the overall allocation is of size at most *D*. Note that |𝒮| = *O*(*K*^*ℓ*^ · *D*^*ℓ*^). We then iterate over all elements of 𝒮 and call LatticeGreedySubroutine with 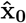 set to each element in 𝒮. We call this entire algorithm *ℓ*-EnumGreedy. Later in this section, we analyze 3-EnumGreedy.

While 3-EnumGreedy runs in polynomial time (specifically, *O*(*K*^4^ · *D*^5^ · *T*_*g*_) time), it is expensive and not practical for large instances. A cheaper algorithm based on LatticeGreedySubroutine computes one solution by starting LatticeGreedySubroutine with **0** as the initial allocation and then computes *K* additional “singleton” solutions by allocating the entire budget to each of the *K* subpopulations. The final solution returned is the best of these *K* + 1 solutions. We call this the SingletonGreedy algorithm. Note that the running time of SingletonGreedy is dominated by LatticeGreedySubroutine.

##### Algorithm 1

LatticeGreedySubroutine 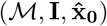

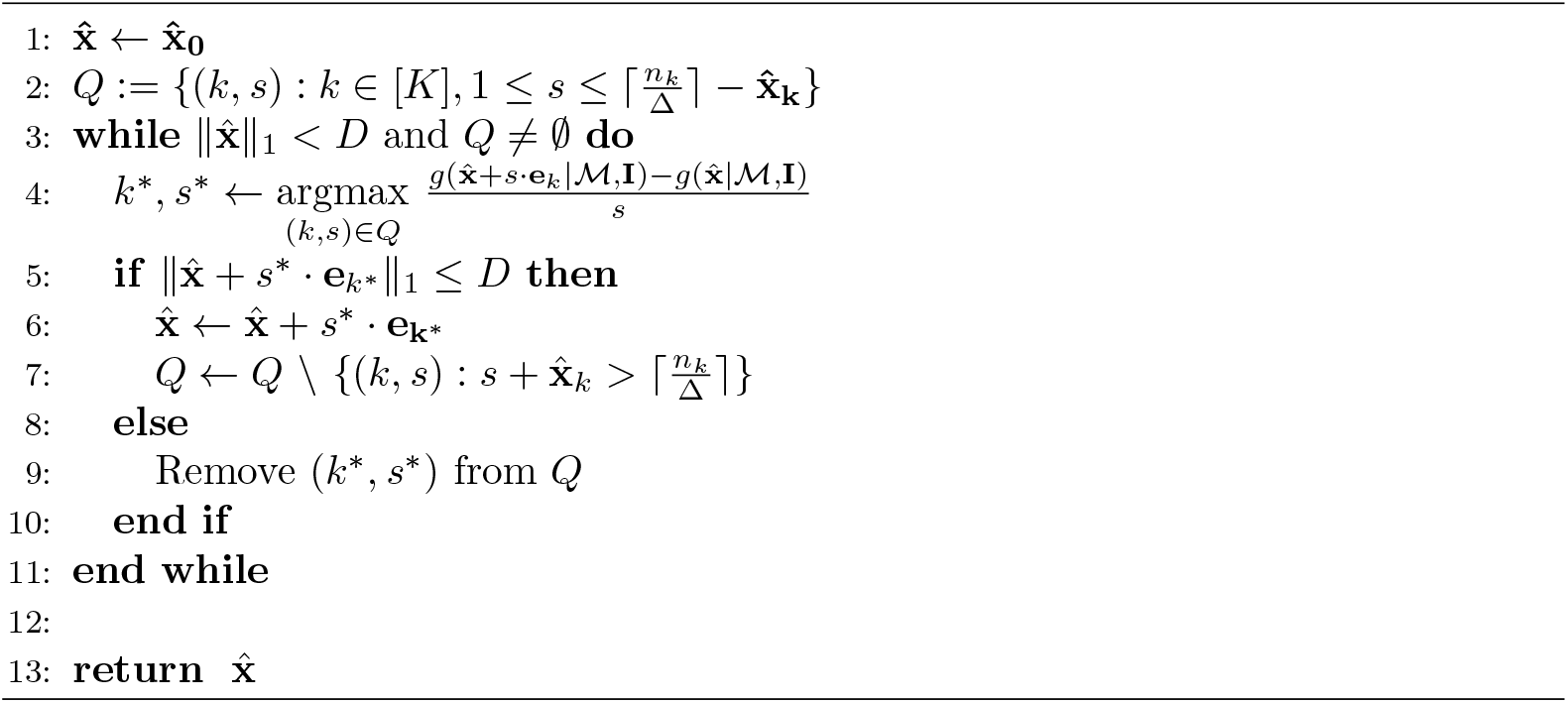

### FastGreedy Description

FastGreedy [25] is a relaxation of LatticeGreedySubroutine that sets an initial threshold and adds the (*k, s*) pair that provides the maximum benefit for each *k* ∈ [*K*] if that benefit exceeds the threshold *τ*_*f*_ of that iteration. In each iteration, the threshold is relaxed for the next round of allocations. *κ*_*f*_ determines the rate at which the threshold *τ*_*f*_ decreases, and *β*_*f*_ approaches the FastGreedy *DR-submodularity ratio* 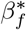 [25], which upper-bounds the DR-submodularity ratio. The rate at which *β*_*f*_ approaches 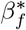 is governed by the parameter *δ*_*f*_. *ε*_*f*_ establishes a lower bound on how small the marginal gain in each iteration can be before the algorithm exits.

FastGreedy is a relaxation of LatticeGreedySubroutine in two ways: **i)** in each iteration, FastGreedy allows allocation to multiple *k* ∈ [*K*], as long as their benefit exceeds an iteration dependent threshold, and **ii)** determines the number of bundles *s* through a binary search subroutine, where LatticeGreedySubroutine exhaustively searches through each (*k, s*) pair.

#### Algorithm 2

FastGreedy (ℳ, **I**, *κ*_*f*_, *d*_*f*_, *ε*_*f*_ ∈ (0, 1))

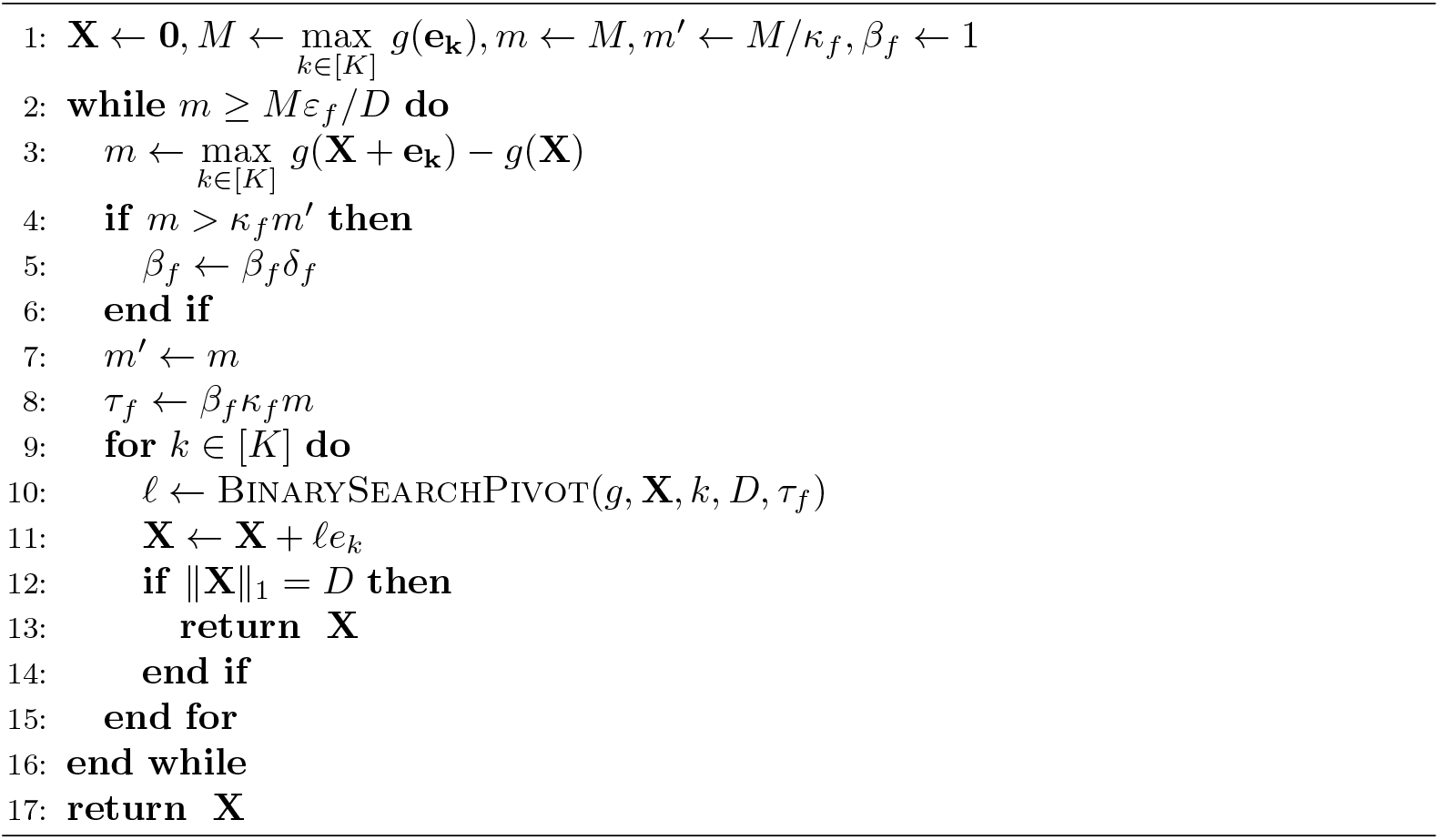

### UnitGreedy Description

On the problem instances we consider, in practice, 3-EnumGreedy, and SingletonGreedy elect to allocate one bundle at a time for a majority of iterations. With this in mind, we consider another more efficient algorithm, UnitGreedy. As shown in the Algorithm 3 pseudocode, UnitGreedy allocates one vaccine bundle to a subpopulation *k* ∈ [*K*], each time selecting a subpopulation that yields the highest marginal gain in the objective function - this is equivalent to converting the lattice into a multiset and running a set greedy algorithm on it (such as the one in [26]). The algorithm continues until the vaccine budget *D* is met. The running time of this algorithm is *O*(*K* · *D* · *T*_*g*_). Note that the marginal gains for the various bundles can be computed (Line 3 in Algorithm 3) in parallel in a straightforward manner, and if we ignore overhead for parallelization, the running time reduces to *O*(*D* · *T*_*g*_).

#### Algorithm 3

UnitGreedy (ℳ, **I**)

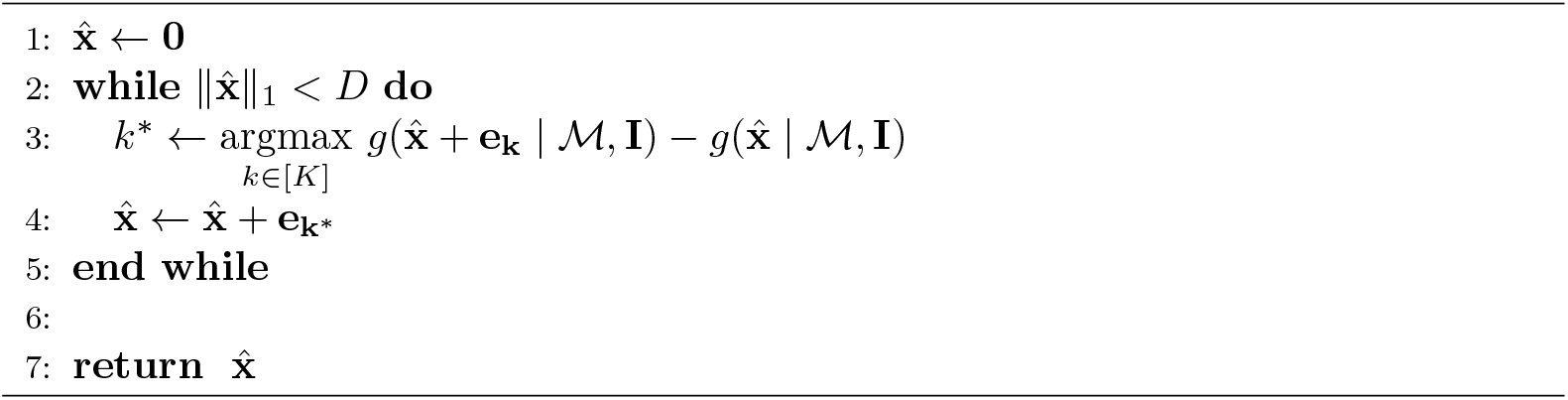

### Approximation guarantees

#### Lattice function submodularity ratios

To analyze the greedy algorithms described above, we utilize the notion of *submodularity ratio* defined in [24]. The submodularity ratio of a function *g* is a quantity between 0 and 1 that is a measure of *g*’s “distance” to submodularity. Since there are two distinct notions of submodularity for lattice functions, as defined in the Background section, there are two associated notions of submodularity ratios, which we now present. To simplify notation, we drop ℳ and **I** and simply use *g*(**x**) for our objective function.

##### Definition 7.

***DR-Submodularity Ratio***. *[24] The DR-submodularity ratio of a function* 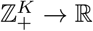 *is defined as*

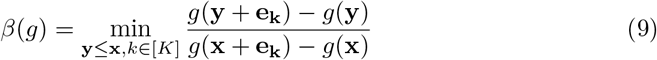

In this definition (and in the next definition below) we designate 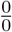 to be 1 and 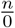 to be ∞ for any positive integer *n*. From this definition it is clear that *β*(*g*) ≤ 1 because **x** = **y** is included in the space that is being minimized over. Furthermore, this definition along with the definition of DR-submodularity (Definition 1) implies that *β*(*g*) = 1 iff *g* is DR-submodular. Thus, the “distance” 1 − *β*(*g*) indicates how far the function *g* is from being DR-submodular. Below we present a similar definition that captures the notion of “distance” of a function *g* from being submodular.

##### Definition 8.

***Submodularity Ratio***. *[24] The submodularity ratio of a function* 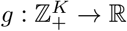*is defined as*

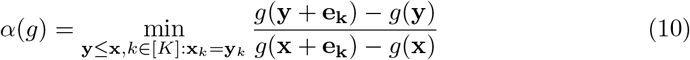

Like *β*(*g*), the submodularity ratio *α*(*g*) also satisfies *α*(*g*) ≤ 1 and 1 − *α*(*g*) indicates how far the function *g* is from being submodular. Since submodularity is a weaker notion than DR-submodularity, an arbitrary lattice function will be “closer” to submodularity than DR-submodularity. Correspondingly, *α*(*g*) ≥ *β*(*g*).

We now present approximation guarantees for 3-EnumGreedy (Theorem 9a), SingletonGreedy (Theorem 9b), and UnitGreedy (Theorem 10). The approximation guarantee associated with FastGreedy can be found in [25].

### Guarantees for 3-EnumGreedy and SingletonGreedy

Theorem 9 provides a guarantee for 3-EnumGreedy and SingletonGreedy. Previously, [20] established approximation guarantees for these algorithms over submodular objective functions, whereas we establish them for more general objective functions.

#### Theorem 9.

*Let* 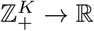 *be an arbitrary monotone function. Let OPT denote the optimal solution to the problem* 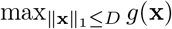.

a. *If* 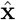 *is the solution returned by 3-*EnumGreedy *then*

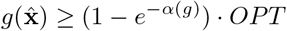
b. *If* 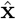 *is the solution returned by* SingletonGreedy *then*

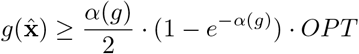

The proof is included in the Supplementary Information.

### Guarantee for UnitGreedy

Here, we provide a version of the approximation guarantee found in [26], which is dependent on the submodularity ratio for set functions [23] and generalized curvature [26]. Their guarantee is applicable to UnitGreedy when we consider the lattice over which we allocate to be a multiset.

#### Theorem 10.

*Let* 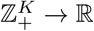 *be an arbitrary monotone function. Let OPT denote the optimal solution to the problem* 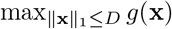.*If* 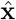 *is the solution returned by* UnitGreedy *then*

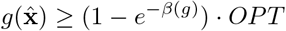

The above results show that the approximation guarantees shown in the literature [20, 28] for greedy algorithms when *g* is a submodular function (on sets or lattices) are more general and apply to arbitrary monotone integer lattice functions. Note that the theorems above also provide a trade-off between approximation-factor and running time. UnitGreedy is the fastest algorithm, but this provides an (1 − *e*^−*β*(*g*)^)-approximation, which is no better than the (1 − *e*^−*α*(*g*)^)-approximation provided by the more expensive algorithm 3-EnumGreedy.

We remark that *ℓ*-EnumGreedy, SingletonGreedy, FastGreedy, and UnitGreedy are well known algorithms for maximizing a submodular function over sets or lattices subject to a cardinality constraint (e.g., [19, 20, 25, 26]). Our main contribution here is to show that 3-EnumGreedy and SingletonGreedy provide approximation guarantees even when the objective function is not submodular and these guarantees degrade gracefully as the objective function becomes less submodular, as measured by the submodularity ratio. We also derive a lattice function based approximation guarantee for UnitGreedy, extending from the set function guarantee provided in [26].

Finally, we note that the POMS algorithm in [24] achieves a max ((1 − *e*^−*β*(*g*)^), *α*(*g*)*/*2 · (1 − *e*^−*α*(*g*)^)) -approximation. Our results show that simple, well-known, and faster greedy algorithms achieve these same approximation factors.

## Experiments

Next, we present a variety of experiments that collectively show that **i)** greedy methods outperform various baseline vaccine allocation algorithms for both MaxCasesAverted and MaxPeaksReduced objectives, **ii)** greedy methods are very close to optimal for all instances for which this comparison was feasible, and **iii)** the greedy methods are considerably faster than POMS [24] (when requiring all algorithms to run until their approximation factors can be guaranteed). We run our experiments at 3 different scales: (i) small-scale experiments: New Hampshire (10 counties, population 1.4 million), (ii) medium-scale experiments: Iowa (99 counties, population 3.2 million), and (iii) large-scale experiments: Texas (254 counties, population 30.03 million). Our code and processed data are part of the Supplementary Information. Experiments were run on AMD EPYC 7763 CPUs with 2 TB RAM.

### Baselines

Our baselines include natural vaccine allocation strategies such as Population, Out-Mobility, In-Mobility, and Random, which assign vaccines to each county proportional to the population, the total mobility originating in the county, the total mobility terminating in the county, and uniformly at random respectively. We also compare our approaches against POMS [24], which works by expanding a random pareto-optimal frontier.

### Data

Our experimental test-beds consist of simulated outbreaks over inter-county mobility graphs for New Hampshire, Iowa and Texas constructed from two separate sources: **(i) FRED** [30] (open source) is a census-based synthetic population contact network, which includes high-resolution social, familial, demographic, and behavioral details, and **(ii) SafeGraph** [31] (open source for academics) provides aggregated and anonymized mobility data from mobile device GPS signals, which provides inferred ‘home’ locations and visits to places of interest (POIs). We derive state-level directed mobility graphs from both data sources, where nodes correspond to counties and directed weighted edges correspond to movement from the source county to the target county.

The mobility graphs constructed using FRED and SafeGraph are similar for New Hampshire and Iowa, except that the SafeGraph mobility graphs have a slightly higher density. For Texas, the density of the FRED mobility graph is an order of magnitude lower than that of SafeGraph. A description of the mobility graph construction and a table of their properties can be found in the Supplementary Information.

### Parameters

We select values of *λ* (infectivity) at approximately 0.347 and 0.535 to result in 20% and 70% of each population becoming infected without vaccination, respectively. We conducted experiments with a wider range of *λ* values (in general, we observed that problem instances with lower values of *λ* are more easily solved by more vaccine allocation methods) and chose two values that represent significantly different levels of infectivity. We performed experiments for New Hampshire, Iowa, and Texas, with a vaccine budget of 10% through 60% of each state’s total population in 10% increments. The parameters *k, n*_*i*_, and *w*_*ij*_ are instantiated according to the data when we constructed the mobility graphs. The parameters *r*_*i*_ scale the infectivity parameter *λ* for each county, and is set in proportion to the population density of each county. We set the initially infected vector **I**_0_ to be 1 for each county. The choice in **I**_0_ does not make a difference in our setting due to the deterministic nature of our model and the small diameter of our mobility graphs (at most 4). *η* and *δ* are set according to [34]. For FastGreedy in New Hampshire and Iowa, we set *κ*_*f*_ = *δ*_*f*_ = 0.96, and in Texas, we set *κ*_*f*_ = *d*_*f*_ = 0.93. For all FastGreedy experiments, we set *ε*_*f*_ = 0. We run each simulation for at least 200 timesteps and terminate the simulation when the disease dies out.

### Performance of greedy methods compared to baselines

In our first experiment, we compare the performance of greedy vaccine allocation algorithms to baseline algorithms using both the FRED and SafeGraph mobility graphs, for both the MaxCasesAverted and MaxPeaksReduced problems. For our small-scale experiment (New Hampshire), we run all four greedy algorithms. For our medium-scale experiment (Iowa), we drop our slowest greedy algorithm 3-EnumGreedy. For our large-scale experiment (Texas), we drop our slowest two greedy algorithms 3-EnumGreedy and SingletonGreedy. For this comparison, we always run POMS for the same amount of time as UnitGreedy. We seek to demonstrate how close the performance of POMS gets to that of UnitGreedy in a simple wall clock time based comparison. We repeat these experiments for six different budgets (expressed as a percentage of the population of the state) for two different values of *λ*. The results for a high infectivity value of *λ* are summarized in Fig 2, 3, and 4. The same experiments for lower infectivity parameter values can be found in the Supplementary Information.

**Fig 2.**
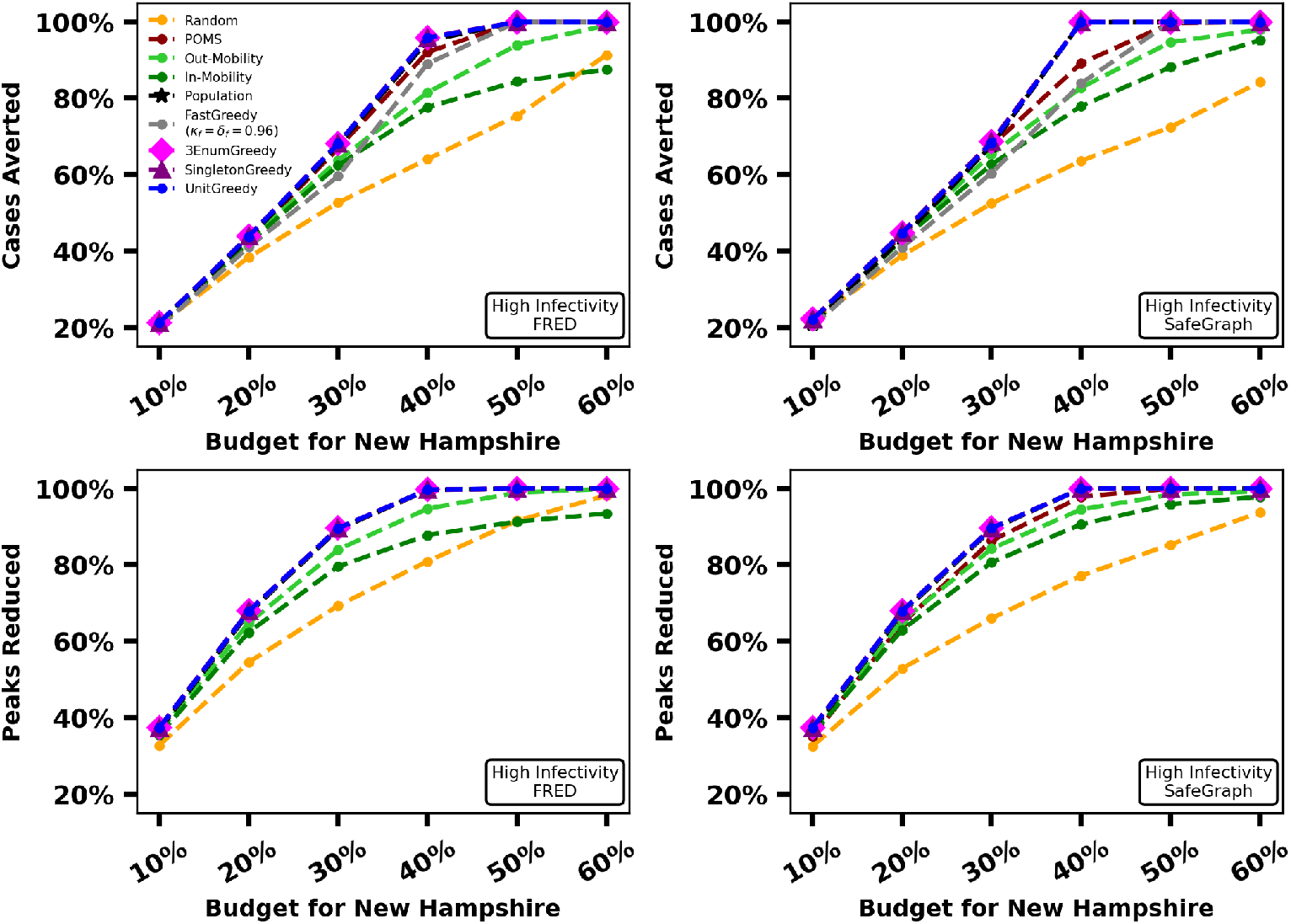
Percentage TotBurden and percentage MaxBurden reduced by all approaches for *λ* = 0.5345 in New Hampshire for FRED (first column) and SafeGraph (second column).

**Fig 3.**
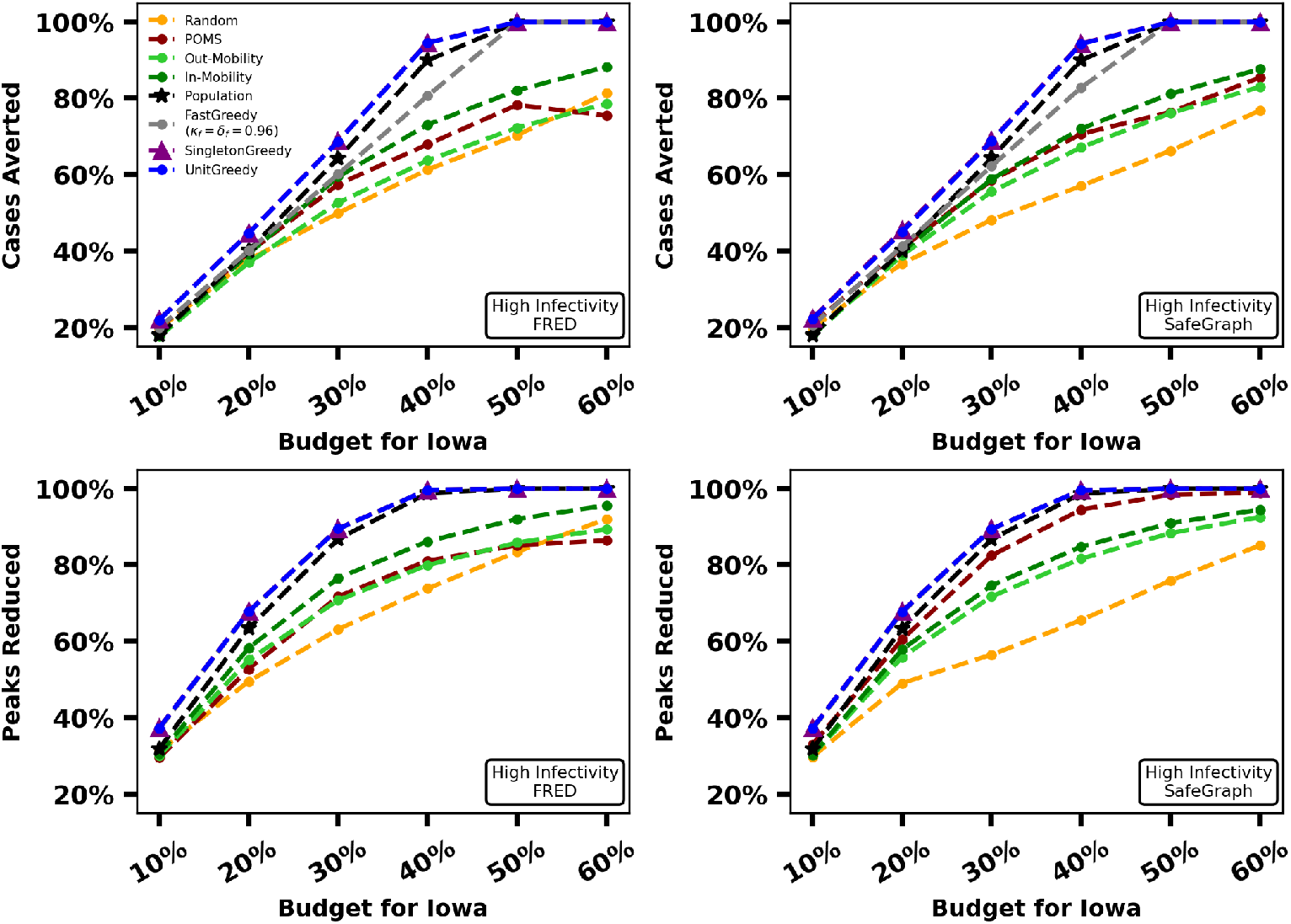
Percentage TotBurden and percentage MaxBurden reduced by UnitGreedy, SingletonGreedy, FastGreedy and baselines for *λ* = 0.535 in Iowa for FRED (first column) and SafeGraph (second column).

**Fig 4.**
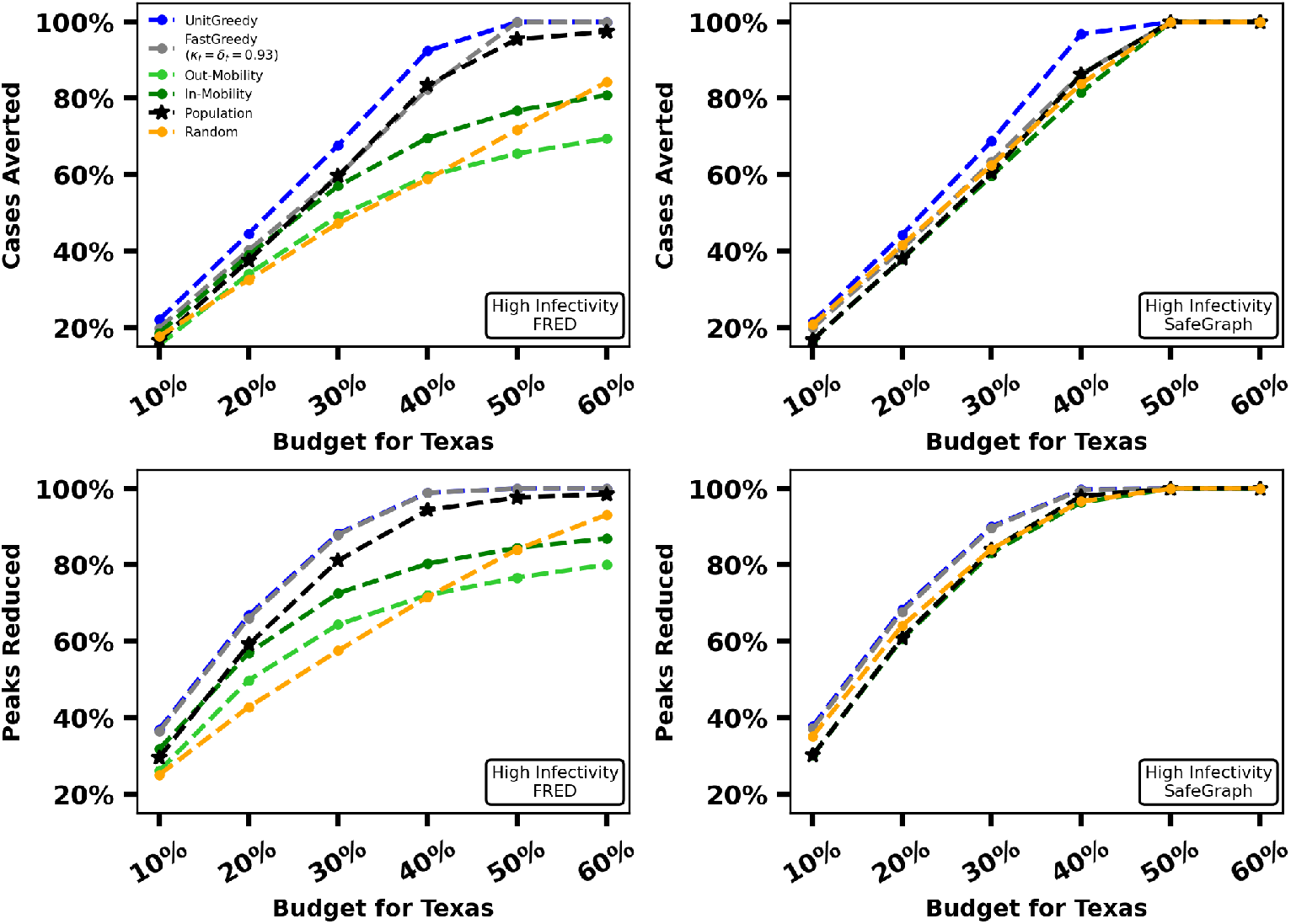
Percentage TotBurden and percentage MaxBurden reduced by UnitGreedy, FastGreedy and baselines for *λ* = 0.525 in Texas for FRED (first column) and SafeGraph (second column).

Fig 2 shows that, for our small-scale experiments, the baselines never outperform the greedy methods. Population and POMS perform on-par with the greedy methods in some instances, particularly in MaxPeaksReduced. We see the performance of baselines relative to the greedy methods decline as the scale of our experiments become larger. As observed in Fig 3, even for our medium-scale experiments, the greedy algorithms outperform each baseline in several settings, while no baseline outperforms the greedy methods. Fig 4 demonstrates that, for our large-scale experiment, UnitGreedy and FastGreedy outperform the baselines by a wider margin than our small and medium-scale experiments over the FRED dataset. This margin is more narrow (with UnitGreedy and FastGreedy still in the lead) over the SafeGraph mobility graph. For SafeGraph, UnitGreedy and FastGreedy perform on-par with the same methods over the FRED data - the difference is primarily in the increased performance of the baselines over SafeGraph. Similar results hold for a lower value of *λ*, which we include in the Supplementary Information. UnitGreedy performs at least on-per with the other greedy methods, all of which employ larger search spaces. In all experiments, after the greedy methods, the Population heuristic performs well, followed by POMS, other baselines, and finally Random. The relatively poor performance of POMS could be attributed to the fact that it requires a long running time to achieve its theoretical guarantee (see Performance-Time Trade-off). The surprisingly good performance of the Population heuristic suggests that it might be a good on-the-field strategy in the absence of mobility data for small problem instances. UnitGreedy substantially outperforms Population and FastGreedy for our large-scale experiment on Texas over FRED data, with TotBurden reduced by up to 8% of the population, which translates to almost 2 million additional cases avoided.

### Near-optimality of greedy algorithms

In this section, we demonstrate that in practice, the greedy algorithms we evaluate return allocations whose objective function value is close to optimal for both MaxCasesAverted and MaxPeaksReduced. Focusing on our small-scale experiment (New Hampshire) using mobility derived from FRED data, we consider 4 problem instances for each of MaxCasesAverted and MaxPeaksReduced, obtained by setting *λ* to 0.347 and 0.5345 and the budget *D* to 2 values (10% and 40% of the population). For these problem instances we compute an optimal solution by exhaustive search and compare the results to that of 3-EnumGreedy, SingletonGreedy, unitgreedy, and fastgreedy

For each problem and problem instance, let OPT denote the objective function value of an optimal solution. Table 3 shows the performance relative to OPT of problem instances for 10% and 40% budgets, high and low infectivity, and both objective functions for each greedy method.

**Table 3.** Approximation factors for each problem instance.

| **NH (FRED)**<br>**10% Budget** | Cases Averted |  | Peaks Reduced |  |
| --- | --- | --- | --- | --- |
| | $\lambda = 0.3475$ | $\lambda = 0.5355$ | $\lambda = 0.3475$ | $\lambda = 0.5355$ |
| 3-ENUMGREEDY | 99.84% | 98.53% | 99.71% | 96.33% |
| SINGLETONGREEDY | 79.02% | 99.04% | 99.71% | 99.33% |
| FASTGREEDY | 79.02% | 95.29% | 92.29% | 95.21% |
| UNITGREEDY | 79.02% | 99.04% | 99.71% | 96.33% |

**Table 3.** Approximation factors for each problem instance
| **NH (FRED)**<br>**40% Budget** | Cases Averted |  | Peaks Reduced |  |
| --- | --- | --- | --- | --- |
| | $\lambda = 0.3475$ | $\lambda = 0.5355$ | $\lambda = 0.3475$ | $\lambda = 0.5355$ |
| 3-ENUMGREEDY | 100% | 99.86% | 100% | 99.97% |
| SINGLETONGREEDY | 100% | 99.86% | 100% | 99.97% |
| FASTGREEDY | 100% | 99.26% | 100% | 99.97% |
| UNITGREEDY | 100% | 99.86% | 100% | 99.97% |

Problem instances for the state of Iowa are much larger and it is not feasible to compute OPT to make a direct comparison. To circumvent this problem, we first note that it is possible to obtain improved versions of Theorems 9(a), 9(b), and 10 by defining “per instance” versions of the DR-submodularity ratio and submodularity ratio. To be specific, let 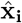 denote the allocation after iteration *i* of UnitGreedy, let **x**^*^ be an optimal solution, and let 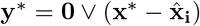.Define

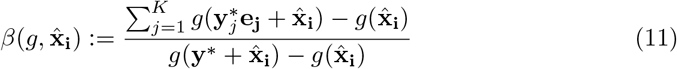

The numerator is the total marginal gain of independently increasing each individual subpopulation’s allocation to the optimal allocation. The denominator is the marginal gain of increasing 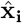 to the optimal solution all at once. If *g* were submodular, it would follow that 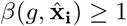, but this guarantee does not hold for an arbitrary *g*(·). It is possible to show that the bound stated in Theorem 10 holds for 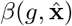, i.e., 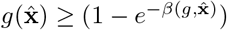 (more on this may be found in the Supplementary Information).

Since we cannot calculate the optimal solution **x**^*^ directly (and 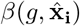) depends on **x**^*^) we cannot calculate 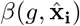 directly either. Instead, we use a sampling method (described in the Supplementary Information) to find an estimate of 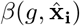, which we denote as 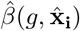. We calculate 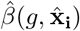 5000 times for each experiment to estimate 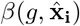.Our key finding is that 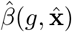 is very close to (or even larger than) 1 for most of our experimental instances, implying that *g* might be close to being submodular in practice. This suggests that the allocation 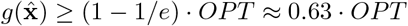.

Estimates for 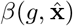 can be found in Table 4. These values indicate worst-case approximation factors for performance on-par (and some exceeding) that of submodular functions for our problem formulations and experimental settings.

**Table 4.** Estimates of 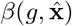 for each problem instance.

| **NH (FRED)**<br>**60% Budget** | Cases Averted |  | Peaks Reduced |  |
| --- | --- | --- | --- | --- |
| | $\lambda = 0.3475$ | $\lambda = 0.5355$ | $\lambda = 0.3475$ | $\lambda = 0.5355$ |
| 3-ENUMGREEDY | 1.03 | 1.01 | 1.74 | 1.02 |
| SINGLETONGREEDY | 1.41 | 1.01 | 1.53 | 1.02 |
| FASTGREEDY | 1.04 | 1.01 | 1.05 | 1.01 |
| UNITGREEDY | 1.51 | 1.02 | 2.66 | 1.12 |
| **IA (FRED)**<br>**60% Budget** | Cases Averted |  | Peaks Reduced |  |
| | $\lambda = 0.3475$ | $\lambda = 0.5355$ | $\lambda = 0.3475$ | $\lambda = 0.5355$ |
| SINGLETONGREEDY | 1.02 | 1.01 | 1.06 | 1.01 |
| FASTGREEDY | 1.04 | 1.01 | 1.05 | 1.01 |
| UNITGREEDY | 1.02 | 1.02 | 1.07 | 1.04 |
| **TX (FRED)**<br>**60% Budget** | Cases Averted |  | Peaks Reduced |  |
| | $\lambda = 0.3475$ | $\lambda = 0.5355$ | $\lambda = 0.3475$ | $\lambda = 0.5355$ |
| FASTGREEDY | 1.12 | 1.03 | 1.04 | 1.03 |
| UNITGREEDY | 1.02 | 1.02 | 1.06 | 1.03 |

### Performance and running-time trade-offs

Here, we compare the performance and running time trade-offs for 3-EnumGreedy, SingletonGreedy, FastGreedy, UnitGreedy and POMS. Let *c*_max_ = max{*n*_*i*_ | *i* ∈ [*K*]}. The approximation guarantee for POMS requires *T* = 2*ec*_max_*D*^2^*K* queries [24]; this makes POMS significantly more expensive to run compared to the greedy methods. The term “query” refers to an evaluation of the objective function *g*(·); here, that evaluation entails running a disease simulation conditioned on a vaccine allocation. Compared to POMS, UnitGreedy requires relatively fewer *T* = *K* · *D* queries. In addition, UnitGreedy is much faster in practice (by Wall Clock Time) than POMS since UnitGreedy is embarrassingly parallel, whereas POMS is much more inherently sequential. These comparisons are presented in Table 5, where we list required iterations and practical run time (extrapolated from 12 hours for POMS).

**Table 5.** FastGreedy, UnitGreedy, SingletonGreedy, 3-EnumGreedy, and POMS comparison with respect to practical running time (estimated for POMS) to achieve approximation guarantee for New Hampshire with 20% and 60% budgets.

| NH (FRED)<br>20% Budget | Queries Required |  | Wall Clock Time |  |
| --- | --- | --- | --- | --- |
| | $\lambda = 0.3475$ | $\lambda = 0.5355$ | $\lambda = 0.3475$ | $\lambda = 0.5355$ |
| FASTGREEDY | 1567 | 70 | 3.9 Minutes | 7.4 Seconds |
| UNITGREEDY | $6.67 \cdot 10^3$ | $6.67 \cdot 10^3$ | 16.7 Minutes | 7.6 Minutes |
| SINGLETONGREEDY | $6.43 \cdot 10^5$ | $7.57 \cdot 10^5$ | 1 Hour | 37.1 Minutes |
| 3-ENUMGREEDY | $6.53 \cdot 10^5$ | $7.58 \cdot 10^5$ | 1 Hour | 37.6 Minutes |
| POMS | $3.63 \cdot 10^{15}$ | $3.63 \cdot 10^{15}$ | $\sim 1.5 \cdot 10^8$ Years | $\sim 1.2 \cdot 10^8$ Years |

| NH (FRED)<br>60% Budget | Queries Required |  | Wall Clock Time |  |
| --- | --- | --- | --- | --- |
| | $\lambda = 0.3475$ | $\lambda = 0.5355$ | $\lambda = 0.3475$ | $\lambda = 0.5355$ |
| FASTGREEDY | 4761 | 2025 | 7.7 Minutes | 3.3 Minutes |
| UNITGREEDY | $2 \cdot 10^4$ | $2 \cdot 10^4$ | 33.2 Minutes | 30 Minutes |
| SINGLETONGREEDY | $3.54 \cdot 10^6$ | $2.7 \cdot 10^6$ | 3.9 Hours | 3.4 Hours |
| 3-ENUMGREEDY | $3.55 \cdot 10^6$ | $2.66 \cdot 10^6$ | 5.1 Hours | 1.5 Hours |
| POMS | $3.27 \cdot 10^{16}$ | $3.27 \cdot 10^{16}$ | $\sim 1.1 \cdot 10^9$ Years | $\sim 1.1 \cdot 10^9$ Years |

FastGreedy introduces an approximation guarantee parameterized by a value which upper bounds the DR-submodularity ratio. Their input parameters can be adjusted to determine the quality required of potential allocation in each iteration, effectively trading performance for speed. When the input parameters to FastGreedy are set so that the performance is maximized, the resulting approximation guarantee is similar to that of UnitGreedy, 3-EnumGreedy, and SingletonGreedy.

## Discussion

Through a combination of theoretical and experimental results, we have shown that even though metapopulation model vaccine allocation problems are inapproximable in the worst case, simple greedy algorithms can be both effective and scalable for these problems.

We provide a possible theoretical explanation for the effectiveness of these greedy algorithms by establishing worst case approximation guarantees in terms of the submodularity ratios of the objective functions of these problems. Specifically, we extend worst case approximation guarantees from the literature for lattice greedy algorithms [20, 25, 26] to the non-submodular objective function setting. Our analysis builds upon prior work on submodular set and lattice function maximization [5, 10, 19, 20, 28, 33]. For specific instantiations of the metapopulation model vaccine allocation problems (e.g., MaxCasesAverted, MaxPeaksReduced) we provide some empirical evidence that the submodularity ratio of the objective functions is high enough (i.e., close enough to 1) to imply that greedy algorithms yield near-optimal solutions to these problems.

The effectiveness of the greedy algorithms we evaluate is maintained across small (New Hampshire), medium (Iowa), and large (Texas) problem scales over two mobility graphs constructed from FRED [30] and SafeGraph [35] data sources. In all problem instances of MVA we evaluate, the greedy methods outperform the baselines, sometimes by quite a significant margin. This difference in performance is typically greatest for a high *λ* (infectivity) value, vaccinating 30% to 50% of the total state’s population for each problem scale. We also demonstrate that the greedy algorithms achieve an approximation factor of over 0.79 for a 10% budget, and an approximation factor of over 0.99 with a 40% budget for both MaxCasesAverted and MaxPeaksReduced problem instances over New Hampshire. Our submodularity ratio estimates for each problem scale approximation guarantees at least match those of submodular objective function maximization.

We observe the performance of the greedy methods are on-par with each other for the Texas FRED and SafeGraph mobility graphs, but the performance of the baselines over the FRED mobility graph are much lower. Because of this, we conjecture that the MVA problem over sparse mobility graphs is harder to solve and we cannot depend on the baselines. Across all experiments, we observe that the MVA problem instances with a lower infectivity value *λ* - infecting approximately 20% of the population - are generally easier to achieve good performance on for all methods.

Moreover, we have parallelized our algorithms to enhance scalability, making the fastest of them take hours to run for the state of Texas. The ability to parallelize the computation allows us to manage the computational demands of large states, ensuring that our methods remain feasible even in high-dimensional datasets. The query complexities for each greedy algorithm (shown in Table 2) further contributes to the feasibility and speed of the fastest two greedy algorithms we present, UnitGreedy and FastGreedy. In addition, it is quite natural to speed up greedy methods by not looking for a locally optimal update in each iteration, but an approximately optimal update, which is a main principle behind the threshold approach of FastGreedy. These features of the greedy methods present a computational advantage with respect to scalability over algorithms such as POMS, introduced in [24].

Despite these contributions, several limitations remain. Our current model is relatively simple and deterministic, assuming homogeneous mixing within populations, which may not capture the complexities of real-world disease spread. Future work could incorporate more sophisticated models, such as agent-based simulations within subpopulations, to better reflect heterogeneous contact patterns. Additionally, the inferred mobility data we use is based on limited sources and does not fully reflect real-world movement patterns, particularly in rural or less structured areas. Expanding to include more comprehensive mobility data, such as transportation networks, would improve accuracy. We also assume preemptive vaccine allocation, which may not be practical in many real-world settings. Addressing non-preemptive vaccine allocation and exploring faster, more scalable algorithms, such as sketch-based methods [25, 36], are promising directions for future research. For this paper, we ran experiments on individual states in isolation without taking physical border effects into account, where in real-world settings, the influence of areas (especially urban) across a state border could have significant impact on vaccine allocation decisions. Additionally, deriving confidence bounds for the estimated submodularity ratios would enhance the robustness of our theoretical guarantees.

## Data Availability

Our experimental framework, all data processing and algorithm code, and output
analysis are available on Zenodo at link https://doi.org/10.5281/zenodo.13882892

## Supporting information

**S1 Fig. Iowa and New Hampshire mobility graphs derived from FRED data**We overlay mobility graphs over maps of Iowa and New Hampshire, where the size of each node is proportional to the population size of the subpopulation in which it is centered. Likewise, the width of each edge *e* ∈ *E*_*ij*_ is proportional to its weight *w*_*ij*_ (number of individuals commuting from subpopulation *i* to subpopulation *j*).

**S2 Fig. Percentage MaxCasesAverted and percentage MaxPeaksReduced for all approaches in New Hampshire under low infectivity**. Most methods are able to save all individuals across all budgets for this small problem instance, with Random being the lowest performing method.

**S3 Fig. Percentage MaxCasesAverted and percentage MaxPeaksReduced for UnitGreedy, SingletonGreedy, FastGreedy and baselines in Iowa under low infectivity**. The effectiveness of the greedy methods is largely unchanged from that of the small problem instances (New Hampshire), but the baseline methods begin to decrease in performance.

**S4 Fig. Percentage MaxCasesAverted and percentage MaxPeaksReduced for UnitGreedy, FastGreedy and baselines in Texas under low infectivity**. For the SafeGraph mobility graph, all methods are able to save most individuals for all budgets, unlike for the FRED mobility graph, where the performance decreases for smaller budgets.

**S1 Text**. Contains Supplementary Information sections A-D, detailing model derivation, approximation guarantee proofs, descriptions of mobility graph construction, parameters, additional experiments, and related work (PDF).

**S1 Table. Comparison of FRED and SafeGraph mobility graph properties**. Contains properties of the mobility graphs constructed from FRED and SafeGraph data in New Hampshire, Iowa, and Texas.

**S2 Table. System specifications for experiments**. Contains information on the CPU type, memory, and storage where we run experiments.

## Acknowledgments

The authors acknowledge feedback from members of the Computational Epidemiology research group at the University of Iowa and the CDC MInD-Healthcare group.

## Author Contributions

**Conceptualization:** Jeffrey Keithley, Bijaya Adhikari, Sriram Pemmaraju.

**Data Curation:** Jeffrey Keithley, Akash Choudhuri.

**Formal Analysis:** Jeffrey Keithley, Bijaya Adhikari, Sriram Pemmaraju.

**Funding Acquisition:** Bijaya Adhikari, Sriram Pemmaraju.

**Investigation:** Jeffrey Keithley, Akash Choudhuri.

**Methodology:** Jeffrey Keithley, Akash Choudhuri, Bijaya Adhikari, Sriram Pemmaraju.

**Project Administration:** Bijaya Adhikari, Sriram Pemmaraju

**Resources:** Bijaya Adhikari, Sriram Pemmaraju.

**Software:** Jeffrey Keithley, Akash Choudhuri.

**Supervision:** Bijaya Adhikari, Sriram Pemmaraju.

**Validation:** Jeffrey Keithley, Sriram Pemmaraju

**Visualization:** Jeffrey Keithley, Bijaya Adhikari.

**Writing – Original Draft:** Jeffrey Keithley, Akash Choudhuri, Bijaya Adhikari, Sriram Pemmaraju.

**Writing – Review & Editing:** Jeffrey Keithley, Akash Choudhuri, Bijaya Adhikari, Sriram Pemmaraju.

